# Predicting Under-five mortality across 21 Low and Middle-Income Countries using Deep Learning Methods

**DOI:** 10.1101/19007583

**Authors:** Adeyinka E Adegbosin, Bela Stantic, Jing Sun

**Affiliations:** School of Medicine, Griffith University, Gold Coast, Queensland; School of Information Communication Technology, Griffith University

## Abstract

**Objectives:** To explore the efficacy of Machine Learning (ML) techniques in predicting under-five mortality in LMICs and to identify significant predictors of under-five mortality (U5M).

**Design:** This is a cross-sectional, proof-of-concept study.

**Settings and participants:** We analysed data from the Demographic and Health Survey (DHS). The data was drawn from 21 Low-and-Middle Income Countries (LMICs) countries (N = 1,048,575). Eligible mothers in each household were asked information about their children and the reproductive care they received during the pregnancy.

**Primary and secondary outcome measures:** The primary outcome measure was under-five mortality; secondary outcome was comparing the efficacy of deep learning algorithms: Deep Neural Network (DNN); Convolution Neural Network (CNN); Hybrid CNN-DNN with Logistic Regression (LR) for the prediction of child survival.

**Results:** We found that duration of breast feeding, household wealth index and the level of maternal education are the most important predictors of under-five mortality. We found that deep learning techniques are superior to LR for the classification of child survival: LR sensitivity = 0.47, specificity = 0.53; DNN sensitivity = 0.69, specificity = 0.83; CNN sensitivity = 0.68, specificity = 0.83; CNN-DNN sensitivity = 0.71, specificity = 0.83.

**Conclusion:** Our findings provide an understanding of interventions that needs to be prioritized, in order to reduce levels of U5M in LMICs. It also demonstrates that deep learning models are more efficacious than a traditional analytical approach.

**Strengths and limitations of this study:**

- The models were tested using a very large data sample, drawn from over 1 million households.
- The survey utilised a cluster sampling approach and are representative of each country included.
- Socio-economic, political and cultural differences between the included countries may limit generalisability of the results.
- The cross-sectional design of the study means we can only infer association and not causality.

## Introduction

Recent global estimates showed that 5.4 million under-five deaths occurred in 2017; this is equivalent to 15,000 deaths every day and 39 deaths per 1000 live births (1). A majority of the children who die before their fifth birthday live in sub-Saharan Africa and South-east Asia; most of these deaths result from preventable and treatable causes (1,2). Although these estimates represent a significant improvement in under-five mortality levels when compared to the levels in the early 1990’s, ‘preventable death of one child is still too many’ (1,2).

High levels of under-five mortality in LMICs is usually a syndromic feature of a weak health system (3), and U5MR is a key barometer of the state of a nation’s health system and an important impact measure that is reliant on health system input such as health financing, health workforce and infrastructure (3,4). These inputs in turn determine health service access, readiness, quality and safety and consequently influences coverage of interventions such as antenatal care coverage, postnatal care, demand for family planning satisfied, skilled birth attendance, care for childhood illnesses, nutritional supplementation etc (4,5).

Studies have shown that improving child survival requires engaging intricately with a host determinants of child health, including biological, environmental and social-economic factors such as level of maternal education, household income, environmental sanitation and hygiene (5–7). The framework of distal and proximate social, environmental and biological determinants was first described by Mosley and colleagues (5). Unfortunately, many developing countries are constrained by limited finances and limited health budgets, and are unable to intervene on all of the determinants of child health at the same time (3). It is therefore increasingly important to identify the most important determinants to be prioritise and to determine the most pressing socio-economic issues that can serve as a starting point for government and policy makers to focus intervention strategy.

Furthermore, intervention measures need to be equity-oriented in order to be effective (9,10). Hence, disaggregated household level monitoring of coverage and impact indicators are crucial for informing policies and programmatic interventions in the sustainable development goal (SDG) era (9). It is important to understand the status of every child as against simply exploring global trends, in order to “leave no one behind” and to “reach the furthest behind first” (10). In light of the SDG pledge, monitoring changes at household or community level may require new ways of engaging with the ‘big data’, that continues to be generated through ongoing household surveys such as the Demographic Health Survey (DHS) and Multiple Indicator Cluster Survey (MICS) (11,12). A shift away from traditional analytical approach may be pertinent and key to effectively monitor health intervention coverage and impact. Machine learning techniques may represent a novel analytical approach to unravel previously unseen trends; these techniques expand on existing statistical approaches and use methods that are not based on a priori assumptions about the distribution of the data” (13).

In a report recently released by the USAID centre for innovation and impact, on the use of Artificial Intelligence (AI) in global health, AI-Enabled population health was identified as one of AI use cases, that could have the greatest impact on improving health quality, cost and access in LMICs (14). AI-Enabled population health encompasses surveillance and prediction, population risk management, intervention selection and intervention targeting (14). In this current study, we explored the efficacy of deep learning as a technique for population health surveillance and intervention targeting.

There have been numerous empirical studies on the various applications of machine learning in hospital settings for prognostication (15,16), triage (17), and prediction of mortality in the hospital setting(18). However, application of machine learning is yet to be demonstrated in population health studies, where it may represent a potential transformative tool (13). The objective of our study is to fill the gap on application of machine learning in population health studies, and other previously highlighted gaps. One of the previously highlighted gaps concerns the need to identify the most important determinants of U5MR. To explore these determinants, we employed a data driven approach by using the random forest algorithm for feature selection, rather than utilising the traditional hierarchical approach for multivariate analysis, which tends to be highly user-driven and usually involving the development of conceptual frameworks that pre-judges the relevance of a limited set of determinants (independent variables) (19). Random Forest is an efficient classification and regression algorithm that combines several randomized decision trees and aggregates their predictions It is especially useful when the number of variables is larger than the number of observations (20).

The RF approach allows an unlimited number of variables or determinants to be incorporated into the model. The algorithm automatically tests several hypothesis and selects features that best predicts the outcome, based on information gained from each variable (16).

Another gap is the need for new ways to gain insights and to unravel previously unseen trends in the prediction of under-five mortality from disaggregated household level data. To fill this gap, we also compared the efficacy of deep learning algorithms: Deep Neural Network (DNN); Convolution Neural Network (CNN); Hybrid CNN-DNN with Logistic Regression (LR) for classifying child survival, and for predicting age of death. Deep learning “discovers intricate structure in large data sets by using backpropagation algorithm to indicate how a machine should change its internal parameter used to compute representation in each layer from the representation in the previous layer” (21). Deep learning algorithms have shown excellent performance in genomics, proteomics, drug discovery, speech recognition, visual recognition, object detection and several other domains (21).

Finally, in this work, we make recommendations on machine learning implementation, and the new regulatory and ethical considerations for the use of novel ML techniques in public health (22).

## Methods

### Data source and analytical tools

We conducted an analysis on DHS data from 21 low and middle-income countries. The DHS is a nationally-representative household survey developed by the United States Agency for International Development (USAID) in the 1980s (23). The survey provides data on fertility, family planning, maternal and child health, gender, HIV/AIDS, Malaria and nutrition (24). In total, over 350 surveys have been carried out in over 90 countries (23). The survey utilizes a two-stage cluster sampling design (24). Combined multi-country data for this study was obtained from the IPUMS-DHS portal (25). Permission to use data for all included countries was granted by the DHS Program. Analysis was conducted using Python software Version 3.7. The programming codes used for the various analysis are accessible on Github using the following link: https://github.com/drulna/u5mr_predict

### Data pre-processing

Any real-world dataset needs pre-processing to convert it into a representation that can be used to train a model. This can heavily affect the model’s performance. This dataset had several irrelevant features, such as IPUMS identifiers created to merge multi-country data. We excluded 14 such features and included 54 features in the final model. However, these records still had missing values. There exist multiple strategies that can be deployed to handle missing values (26). We used the “Forward Fill” strategy in which every missing value is replaced by next real values for each column. This clean and pre-processed data was used for the rest of the analysis (26).

### Feature selection

We use random forest to check feature importance with respect to its predictive power. Figure 2 shows the feature importance (red bar) and variance of each tree in random forest (black vertical line). It can be observed that “Duration of breast feeding” has the most importance to predict a child’s death. However, there are some features that are of limited importance. We perform feature selection based on this information. We drop all features whose importance are less than 0.001, because we found that the accuracy of the classifier does not improve beyond this level, and adding the additional attributes only creates unnecessary additional computational overhead. In total, 34 features fell within our cut-off for feature importance and included in the final model. For comparing the utility of feature selection, we perform two experiments. One without feature selection (on all original 54 features) and one with feature selection (on selected 34 features).

**Figure 1:**
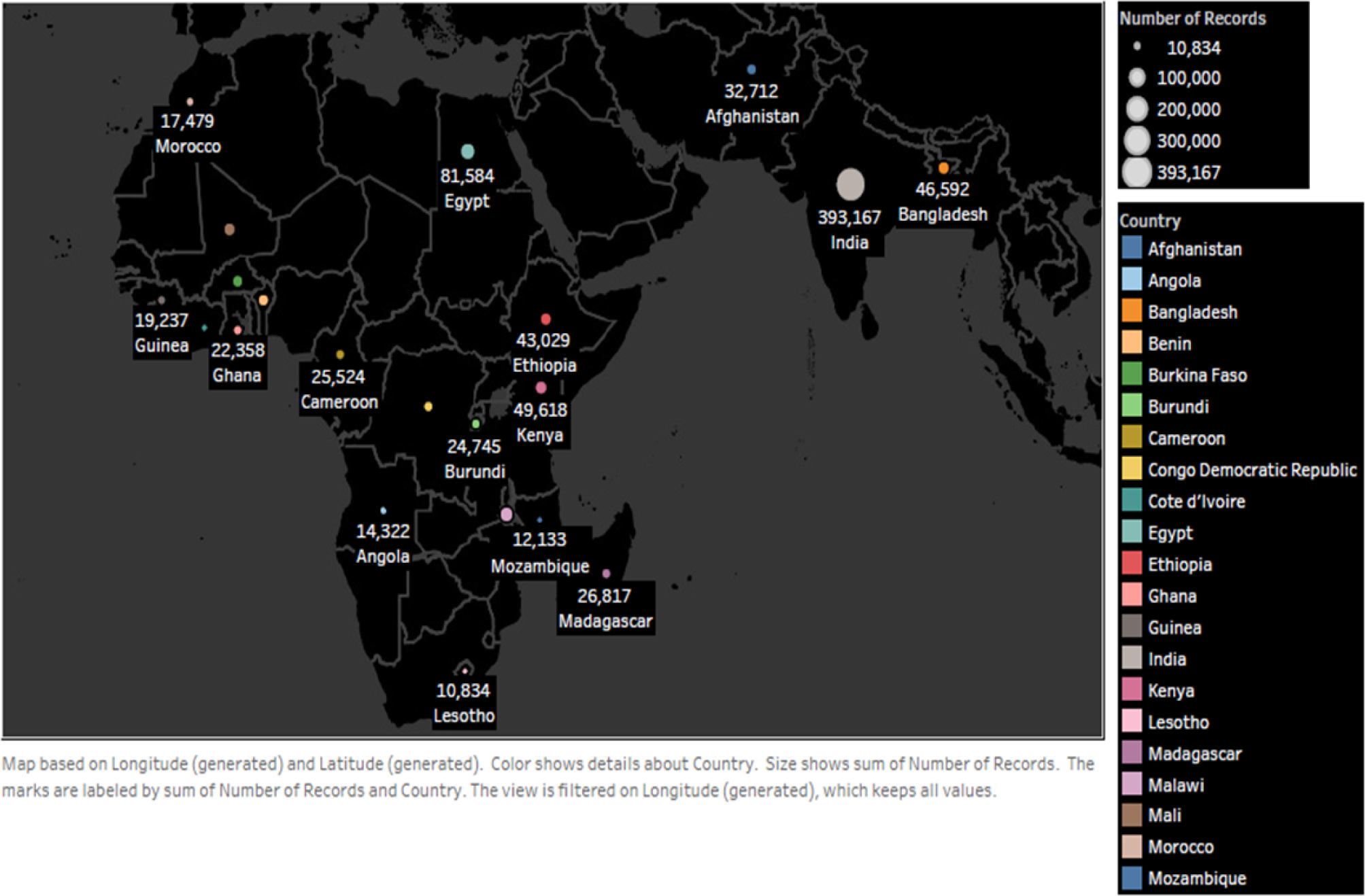
Total sample population per country.

**Figure 2:**
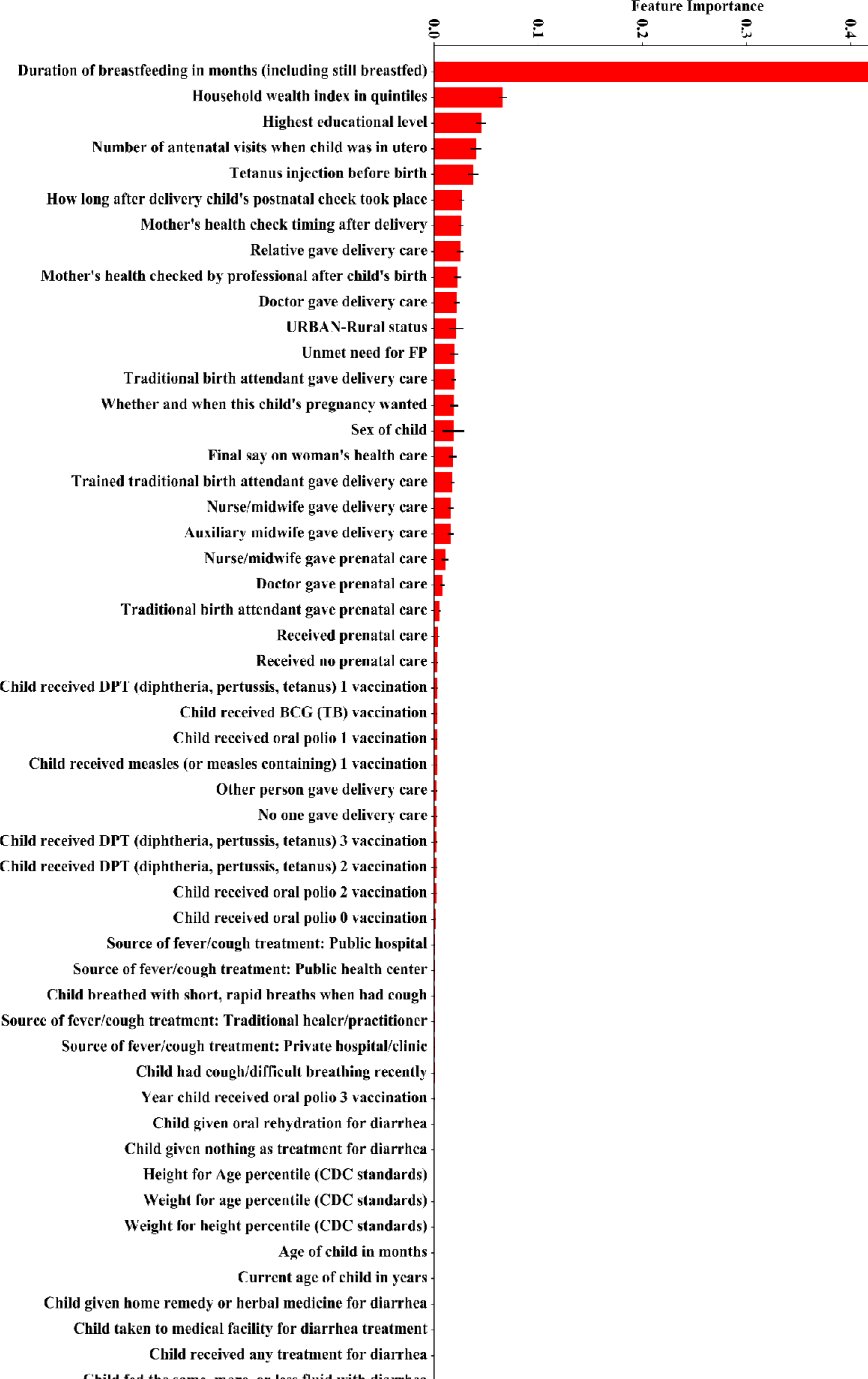
Feature Importance using Random Forest.

### Model Selection

We selected multivariate logistic regression (LR) as an example of traditional model (16). Three deep learning techniques (DNN, CNN and DNN-CNN) were selected as modern machine learning approaches. For all the four models, we pose this problem as a multi-class problem, such that each value in the label is assigned an integer and then we binarize the output (i.e. one-hot encoding). All categorical attributes are also converted to numerical i.e dummy variables, by mapping each unique value to a number. After careful consideration we concluded that the best ratio for training is 75% of the data, while the remaining 25% of the data is reserved for testing purposes. This choice is in line with literature and close to 80/20, which is quite a commonly utilized training/testing ratio, often referred to as the Pareto principle. We compare the performance of logistic regression (LR) as a representative of traditional model, with three deep learning methods: deep network (DN), convolutional neural network (CNN), and hybrid CNN-DN network.

### Model Evaluation

We evaluated the performance of each model using a ROC plot, we also derived the weighted precision, sensitivity (also known as recall), specificity, f1-score, and area under the curve (AUC) for each model. The formula for calculating the performance metrics are as follows:

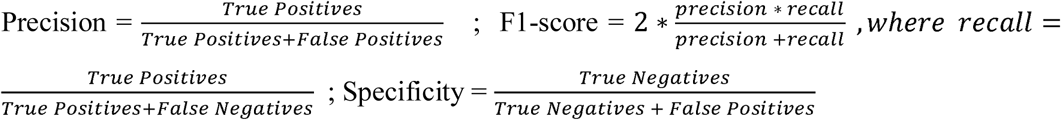

The models were evaluated before and after feature selection. Analysis was initially conducted using all pre-selected variables. We thereafter optimized the various models based on empirical results from the random forest analysis. As this is a multi-class problem, the ROC plots and performance metrics are all based on micro-averages.

## Results

### Characteristics of the study population

A total population size of (N = 1, 048,575) was included in the study, the sample was drawn across 21 LMICs countries. The sample size in each of the included countries is shown in figure 1. The mean age of the total population is 1.89 (± 1.40). Majority (N = 1,100,211; 72.7 %) resides in rural areas. Just under half (N = 636,882; 45.2 %) were in the lowest two wealth quintile (Q1 and Q2). Majority (N = 1,100,262; 73.2%) were uneducated or had only primary education (See Table 1.)

**Table 1:**
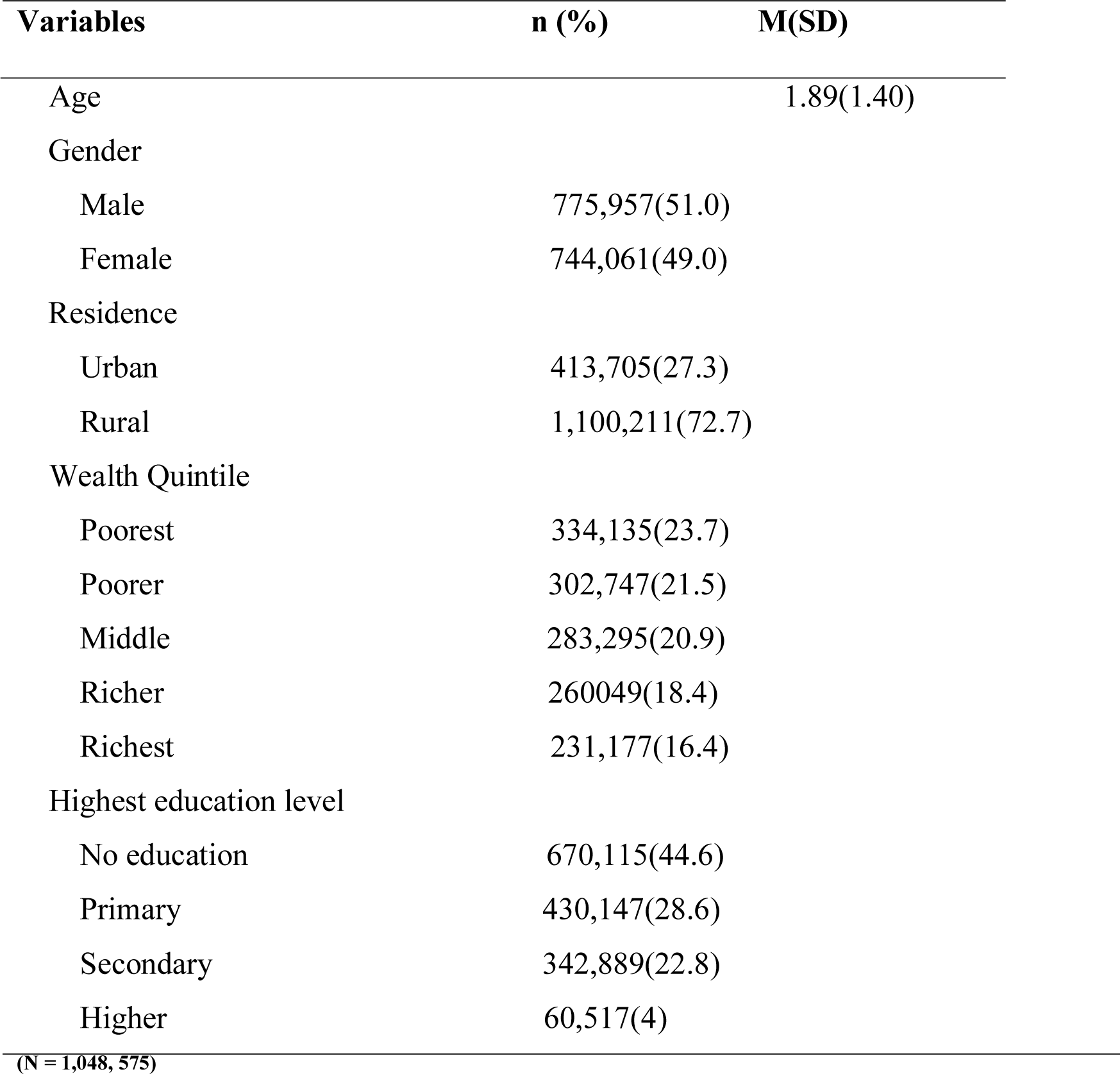
Descriptive analysis of the study population

| Variables | n (%) | M(SD) |
| --- | --- | --- |
| Age |  | 1.89(1.40) |
| Gender |  |  |
| Male | 775,957(51.0) |  |
| Female | 744,061(49.0) |  |
| Residence |  |  |
| Urban | 413,705(27.3) |  |
| Rural | 1,100,211(72.7) |  |
| Wealth Quintile |  |  |
| Poorest | 334,135(23.7) |  |
| Poorer | 302,747(21.5) |  |
| Middle | 283,295(20.9) |  |
| Richer | 260,049(18.4) |  |
| Richest | 231,177(16.4) |  |
| Highest education level |  |  |
| No education | 670,115(44.6) |  |
| Primary | 430,147(28.6) |  |
| Secondary | 342,889(22.8) |  |
| Higher | 60,517(4) |  |

(N = 1,048, 575)

### Feature Importance

We found that the most important determinants of U5MR are duration of Breast feeding, household wealth index and maternal education level. Other key determinants that were identified includes: the number of antenatal visits when the child was in-utero and content of antenatal care Tetanus immunization. (See Fig. 2)

### Model Comparisons (Before Feature Selection)

Comparison of the performance of the models before feature selection reveals that hybrid of CNN-DN performs the best in terms of all metrics (sensitivity = 0.68, specificity = 0.83), while LR performs the worst (sensitivity = 0.47, specificity = 0.53). (see Table 2).

**Table 2:**
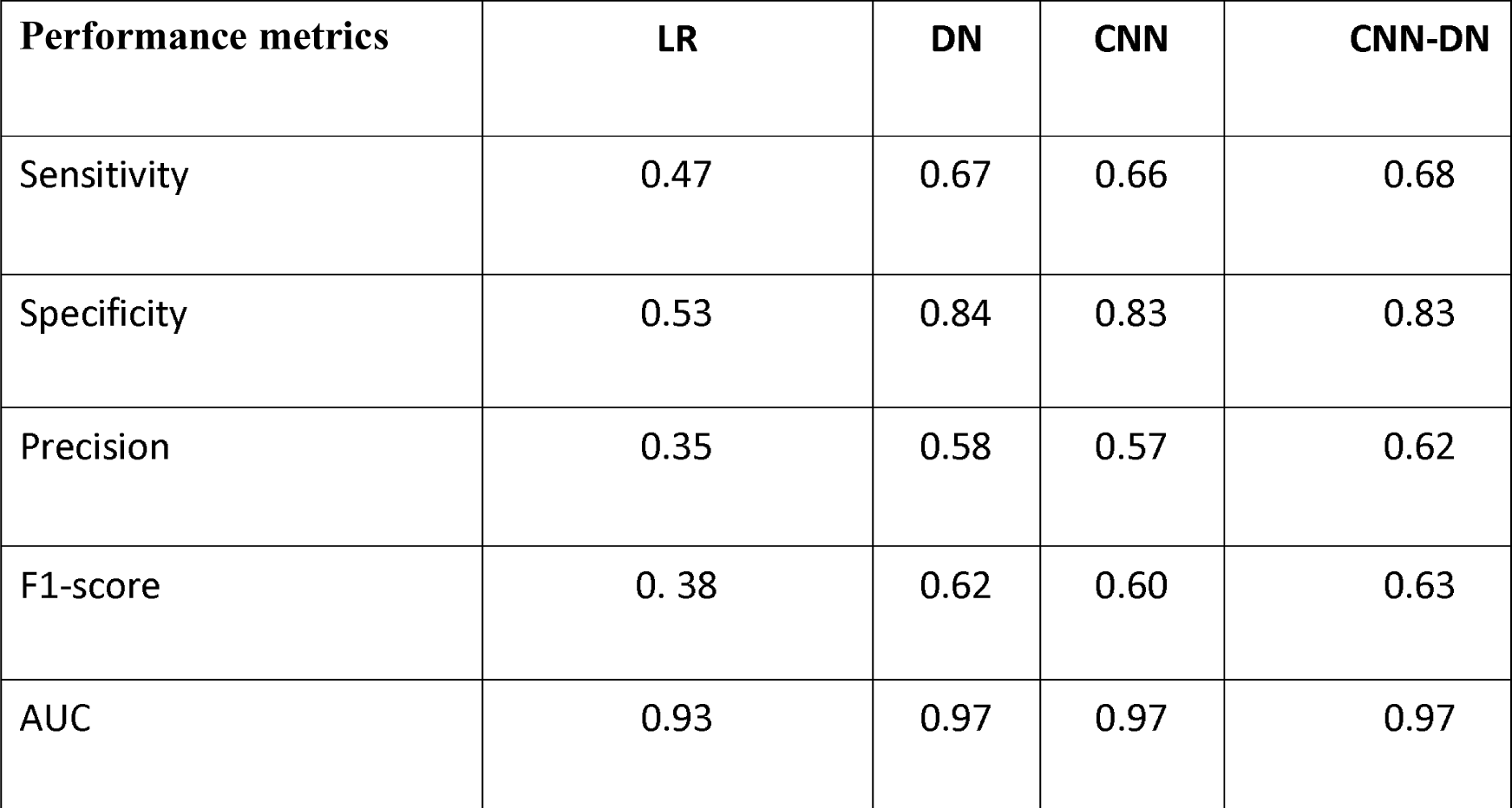
Performance Comparison: (Without Feature Selection)

| Performance metrics | LR | DN | CNN | CNN-DN |
| --- | --- | --- | --- | --- |
| Sensitivity | 0.47 | 0.67 | 0.66 | 0.68 |
| Specificity | 0.53 | 0.84 | 0.83 | 0.83 |
| Precision | 0.35 | 0.58 | 0.57 | 0.62 |
| F1-score | 0. 38 | 0.62 | 0.60 | 0.63 |
| AUC | 0.93 | 0.97 | 0.97 | 0.97 |

Figure 3 shows the ROC curves for all the classifiers. It shows that hybrid CNN-DN model outperforms all other models.

**Figure 3:**
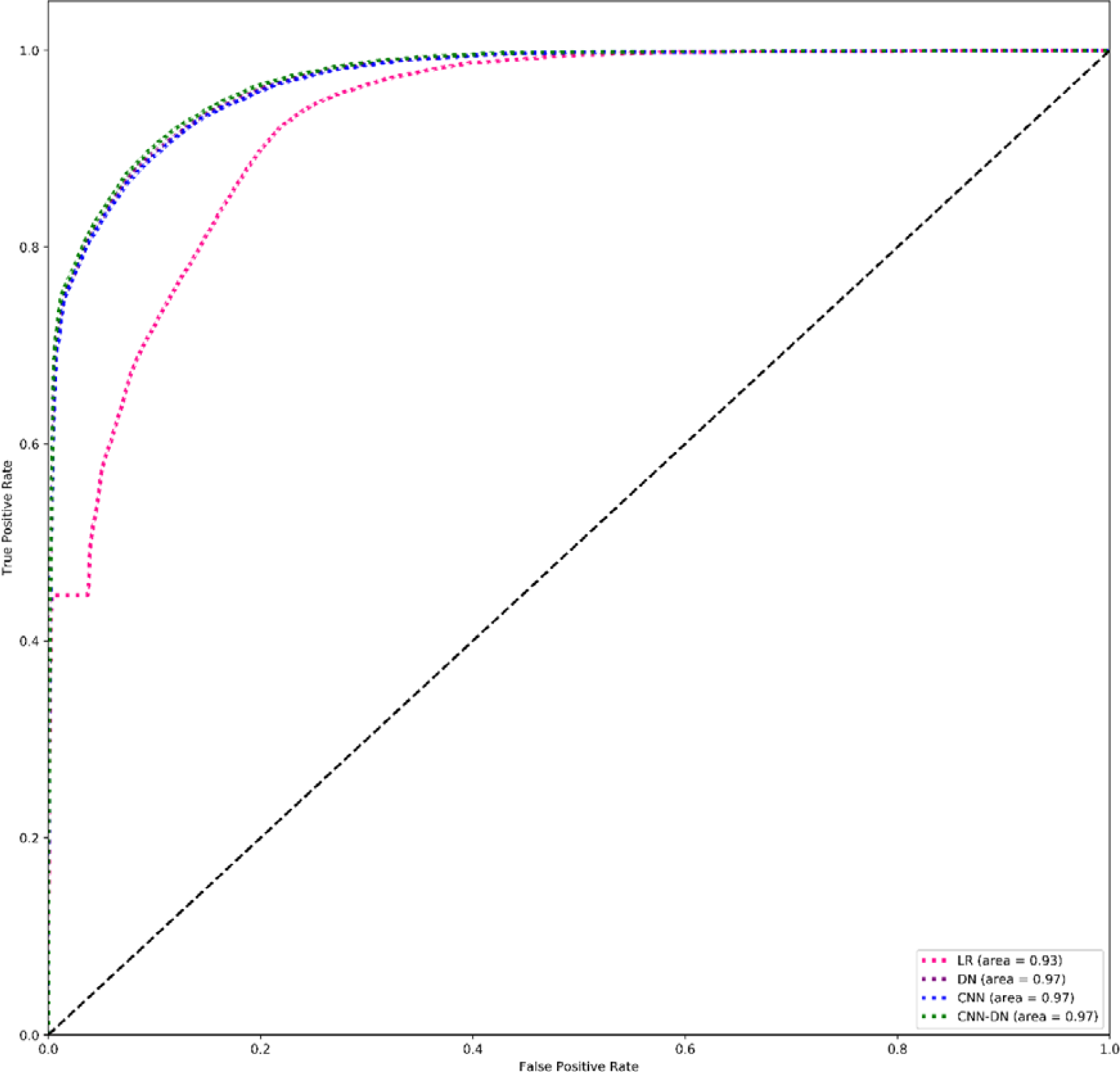
Micro Average ROC Curve (Before Feature Selection)

### Model Comparisons (After Feature Selection)

We found that feature selection does not improve the performance of logistic regression. However, for all deep learning-based models, feature selection results in performance gain. The most performance gain is shown by CNN-DN, (sensitivity = 0.71, specificity = 0.83). CNN-DN model performs the best out of all classifiers in both settings i.e. before feature selection and after feature selection. (see Table 3)

**Table 3:** Metrics comparison after feature selection

| Performance metrics | LR | DN | CNN | CNN-DN |
| --- | --- | --- | --- | --- |
| Sensitivity | 0.47 | 0.69 | 0.68 | 0.71 |
| Specificity | 0.53 | 0.83 | 0.83 | 0.83 |
| Precision | 0.35 | 0.62 | 0.62 | 0.67 |
| F1-score | 0.38 | 0.63 | 0.62 | 0.67 |
| AUC | 0.93 | 0.97 | 0.97 | 0.97 |

In figure 4, we present ROC curves for all the classifiers. It shows that hybrid CNN-DN model remains the top performer of all the models.

**Figure 4:**
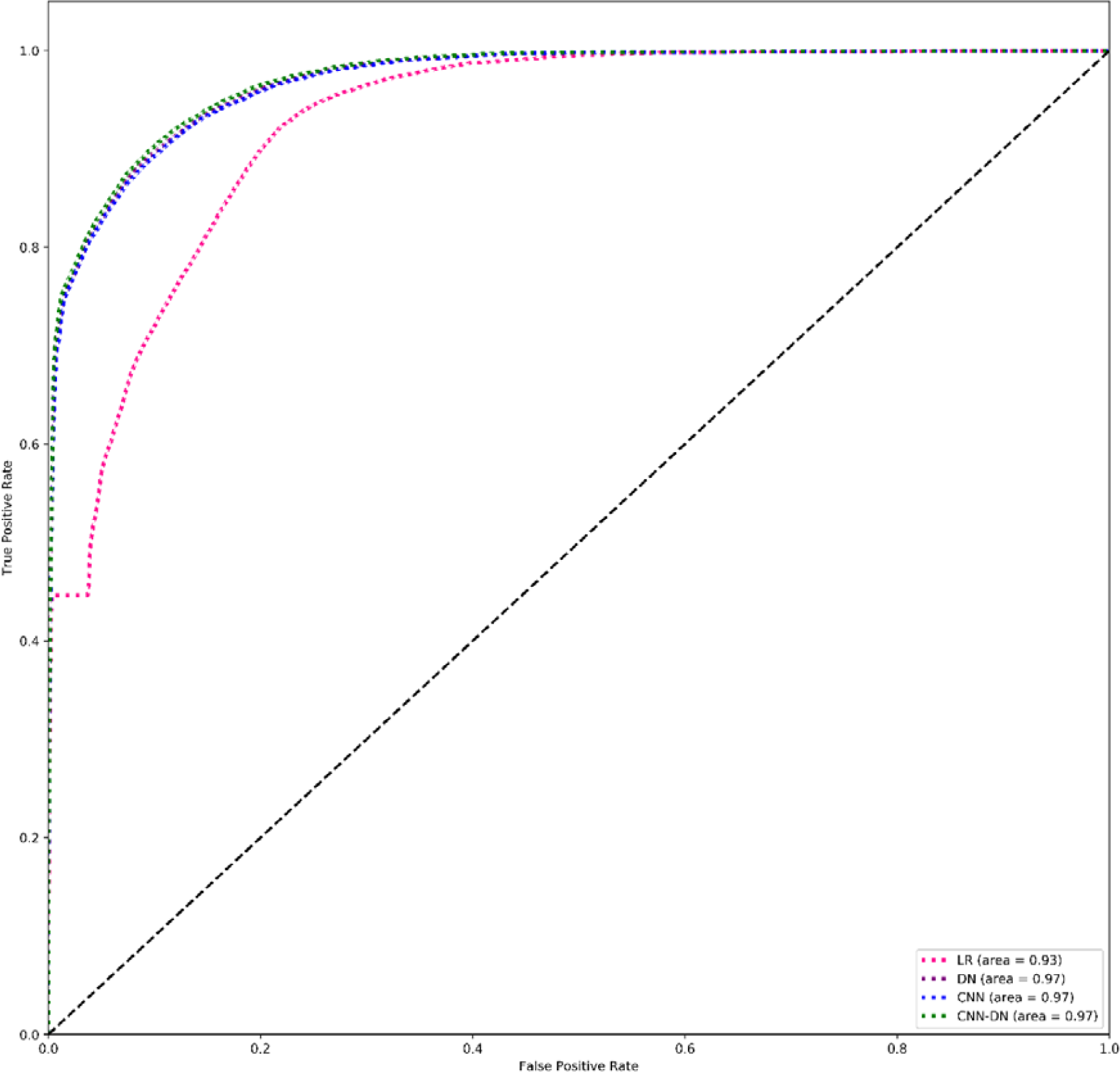
Micro Average ROC after Feature Selection.

## Discussion

We found that the most important determinants of U5M are duration of breast feeding, household wealth index and maternal education level. Previous studies corroborate our findings. It has been shown that children breastfed for a longer duration have lower infectious disease morbidity and mortality, and better chance of survival than those who are breastfed for shorter periods, or not breastfed at all (27). Multiple studies have also shown that early initiation of breast feeding, and exclusive breast feeding reduces both neonatal and early infant mortality (27,28). It has been estimated that scaling up breast feeding can help prevent an estimated 823,000 child deaths across 75 high-mortality LMICs (27). However, just over a third of the total children population in low- and middle-income countries are breastfed; these levels are even lower in industrialized nations (27). Effective approach to scaling up breast feeding practices have been discussed in past literature For example, the Alive and Thrive initiative in rural Burkina Faso involved the utilisation of a multidimensional approach, which combines interpersonal communication and community mobilisation activities to improve breast feeding knowledge, beliefs and skills; this invariably improved breast feeding outcomes (29). A similar approach may be adopted and tailored to the needs in other countries, to scale up breast feeding practices globally. In addition to breast feeding, household wealth index and level of maternal education was also identified as important determinants of child survival in our study. We found that the household wealth index was a slightly more important determinant compared to level of education. This finding however contradicts the work of Fuchs and colleagues, where they argued that mother’s education is the fundamental determinant of child mortality and is relatively more important than income level. They argued that education impacts the child’s health through better maternal health, increased health-specific knowledge, avoidance of traditional, harmful behaviours, greater economic resource as a consequence of education and general female empowerment (29). They however highlighted that other social scientists have often considered education and income as generally highly correlated and tend to be regarded as interchangeable indicators of socioeconomic status (29). Other key determinants that we identified are the number of antenatal visits when the child was in-utero and content of antenatal care (Tetanus immunization), post-natal care, place of residence (urban-rural status), family planning, desirability of the child and sex of the child. These determinants are congruent with the multivariate model of proximal and distal determinants of U5M that have been previously described in literature (5).

Our findings regarding the superiority of machine learning over traditional approaches such as logistic regression in predictive analysis are also in line with findings elsewhere (16,30).

This study however has some limitations. Firstly, this is proof of a concept cross sectional study; hence, we can only draw inference on associations, and not on causality. Secondly, we did not measure change over time. Future studies should consider incorporating temporal data points, in order to draw inference on changes over time, and possibly causality. Finally, we did not explore individual country, regional and subgroup level variations and cannot conclude that the degree of association is the same across different countries and sub-groups, due to differences in socio-economic, geographical, cultural and political realities. Hence, future studies should consider disaggregating with stratifiers such as income, education, and place of residence, in order to explore sub-group differences.

### Recommendations for ML Implementation, governance and ethics

Our recommendations regarding the implementation and regulation of machine learning are in four folds. Firstly, there is a burgeoning risk that the adoption and benefits of ML may be imbalanced (31). High income countries are beginning to increasingly adopt and benefit from deploying some of these novel technologies; therefore, there is the risk of extending the disparity between poor and rich countries even further. To achieve equity in the implementation of this technology, there is a need for capacity building across board and collaborative use of technological resources between low- and high-income countries.

Secondly, regarding AI research governance and ethics (regulation), the capabilities of AI application in public health are not yet fully understood, and its application is still evolving. This implies that any regulatory attempt will effectively require understanding the capabilities of AI as a tool in public health and medicine. Similar to other medical research endeavours, the regulatory framework and ethical guidelines will have to evolve, as our understanding of the application of AI evolves. As such, we posit that there is a concordance between regulation, governance, research and development of AI technology. In the light of this, we suggest collaboration between research institutions, academic stakeholders, policy makers, and regulatory authorities. There is a need to engage with all stakeholders across the spectrum of AI research, development and ethics.

Thirdly, we believe that existing medical research ethical guidelines are highly applicable and cover several aspects of ML research. However, there is a need to strengthen regulatory aspects pertaining to data security and protection. The growth in the adoption of ML analytical techniques will usher an increase in the level of data transactions and with this, comes the potential risk of breaches to health data privacy. There are existing capabilities to re-identify anonymised data, using a few parameters within the data. Hence, regulatory efforts need to focus on data security, especially reducing the risks of data re-identification.

Fourthly, as knowledge and application of AI continues to grow in leaps and bounds, and while regulatory efforts are still rudimentary and trying to catch up, we envisage a vacuum in governance, that will have to be filled. As such, there may be a need for the development and ratification of regulatory framework, which may be possible through the collaboration of multiple stakeholders.

## Data Availability

Data may be accessible through permission from the DHS program

htttps://www.dhsprogram.com

## Conclusions

This study demonstrates the superiority of machine learning as a tool for understanding previously unseen insights in large global health data. We have shown that machine learning algorithms such as random forest, may be more insightful than the user dependent traditional hierarchical approach of testing a limited set of determinants for outcome prediction in multivariate analysis. Using random forest, we found that duration of breast feeding, household wealth index, and level of maternal education are the most important determinants of U5MR. In addition, we also show that deep learning algorithms are more sensitive and specific for the prediction of U5MR and this finding may be applicable to other multivariate models, for data-rich population studies.

Going forward, the most important implication of this study is that if deep learning algorithms such as the one we describe in this study, are deployed in production in combination with spatial data, it is possible to identify and flag children who are most at risk and not likely to survive until the age of five, such that necessary interventions can be targeted to communities where those children live. To the best of our knowledge, there are no existing studies that have investigated under five mortality, using a similar analytical approach.

## Author contributions

AEA conceptualized the study, conducted the data extraction, analysed the data and wrote the first draft of the manuscript. JS and BS contributed to the conceptualization of the study and proofread the document.

## Availability of data and material

The datasets generated and analysed during the current study are available subject to permission from the DHS program, in the [IPUMS-DHS] repository, [https://www.idhsdata.org/idhs/index.shtml].

